# Does Lockdown Decrease the Protective Role of Ultraviolet-B (UVB) Radiation in Reducing COVID-19 Deaths?

**DOI:** 10.1101/2020.06.30.20143586

**Authors:** Rahul Kalippurayil Moozhipurath, Lennart Kraft

## Abstract

**Background:** Nations are imposing unprecedented measures at large-scale to contain the spread of COVID-19 pandemic. Recent studies indicate that measures such as lockdowns may have slowed down the growth of COVID-19. However, in addition to substantial economic and social costs, these measures also limit the exposure to Ultraviolet-B radiation (UVB). Emerging observational evidence indicate the protective role of UVB and vitamin D in reducing the severity and mortality of COVID-19 deaths. In this observational study, we empirically outline the independent protective roles of lockdown and UVB exposure as measured by ultraviolet index (UVI), whilst also examining whether the severity of lockdown is associated with a reduction in the protective role.

**Methods:** We apply a log-linear fixed-effects model to a panel dataset of 162 countries over a period of 108 days (n=6049). We use the cumulative number of COVID-19 deaths as the dependent variable and isolate the mitigating influence of lockdown severity on the association between UVI and growth-rates of COVID-19 deaths from time-constant country-specific and time-varying country-specific potentially confounding factors.

**Findings:** After controlling for time-constant and time-varying factors, we find that a unit increase in UVI and lockdown severity are independently associated with 17% [-1.8 percentage points] and 77% [-7.9 percentage points] decline in COVID-19 deaths growth rate, indicating their respective protective roles. However, the widely utilized and least severe lockdown (recommendation to not leave the house) already fully mitigates the protective role of UVI by 95% [1.8 percentage points] indicating its downside.

**Interpretation:** We find that lockdown severity and UVI are independently associated with a slowdown in the daily growth rates of cumulative COVID-19 deaths. However, we find consistent evidence that increase in lockdown severity is associated with a significant reduction in the protective role of UVI in reducing COVID-19 deaths. Our results suggest that lockdowns in conjunction with adequate exposure to UVB radiation might have provided even more substantial health benefits, than lockdowns alone. For example, we estimate that there would be 21% fewer deaths on average with sufficient UVB exposure while people were recommended not to leave their house. Therefore, our study outlines the importance of considering UVB exposure, especially while implementing lockdowns and may support policy decision making in countries imposing such measures.

## 1 Introduction

Nations are imposing unprecedented non-pharmaceutical intervention measures at large-scale to contain the spread of COVID-19 pandemic^1^. Recent studies indicate that non-pharmaceutical intervention such as lockdowns, ceasing business operations and school closures substantially may have slowed down the growth of COVID-19^1–3^, indicating its protective role. Emerging observational evidence on the epidemiology of COVID-19 indicate that vitamin D deficiency might be a risk factor for COVID-19 deaths^4–7^ and also indicate the protective role of the significant source of vitamin D - Ultraviolet-B radiation (UVB)^8^ in reducing COVID-19 deaths. In addition to substantial economic and social costs, an unintended consequence of lockdown is the likelihood of limited exposure to UVB. However, to the best of our knowledge, no empirical study has explored the association between the severity of lockdown, the subsequent reduction in UVB exposure and the number of deaths attributed to COVID-19 (COVID-19 deaths).

In this observational study, we empirically outline the independent protective roles of lockdown and UVB as measured by ultraviolet index (UVI) and subsequently examine, whether the severity of lockdown is associated with a reduction in the protective role of UVB. After controlling for time-constant and time-varying factors, we find that a unit increase in UVI and lockdown severity are independently associated with 1.8 percentage points (p.p) and 7.9 p.p decline in COVID-19 deaths growth rate representing a decline of 17% and 77%, respectively. These declines indicate the protective roles of UVI and lockdowns. Surprisingly, the widely utilized lockdown with least severity and (e.g., recommendation to not leave the house) already practically fully mitigates the protective role of UVI by 1.8 p.p which represents a decline of 95%.

## 2 Association of Lockdown Severity & UVB Radiation with COVID-19 Deaths

In general, non-pharmaceutical interventions such as the lockdowns aim to reduce the likelihood of transmission of the virus by limiting the movement of people, reducing the contact among individuals via restricting economic activities such as closing restaurants^2^. Some early studies indicate that such large-scale measures might have slowed down the growth rate of COVID-19 infections indicating its protective role, thereby providing direct health benefits^2,3,9^. However, the other indirect health consequences (e.g., reduced exposure to UVB radiation) of such policies are largely unknown.

In addition to substantial economic and social costs of lockdowns, an unintended health consequence of such measures may be the likelihood of limited exposure to UVB radiation. Prior studies indicate that UVB radiation plays a protective role in human health^10–14^. Humans get vitamin D via either diet (natural food, fortified food or supplements) or skin synthesis by UVB radiation exposure ^15^. Likelihood of UVB exposure and subsequent vitamin D synthesis undergo substantial variation according to several time-varying and time-constant factors such as latitude^15^, seasons^15^, time of the day^15^, lifestyle^16,17^, mobility^18^, age^15^, skin pigmentation^15^ and obesity^19^. Prior studies associate vitamin D deficiency with the likelihood of weakened immune response ^20–22^, infectious respiratory diseases ^15,23,24^ and the severity and mortality ^25^.

In Figure 1, we summarize these different factors explaining the potential association between the severity of lockdown and subsequent reduction in the likelihood of UVB exposure with the COVID-19 deaths. Early evidence indicates that lockdown severity^1^ as well as weather factors such as temperature and humidity may reduce the likelihood of transmission of SARS-CoV-2 virus which causes COVID-19^26^. Even though UV radiation may help in reducing the likelihood of transmission by inactivating viruses in fomite transmission^27^, emerging epidemiological evidence related to COVID-19 suggests a protective role of UVB and the plausible role of vitamin D in improving immunity and decreasing the likelihood of COVID-19 severity and mortality^4–8,28^.

**Figure 1:**
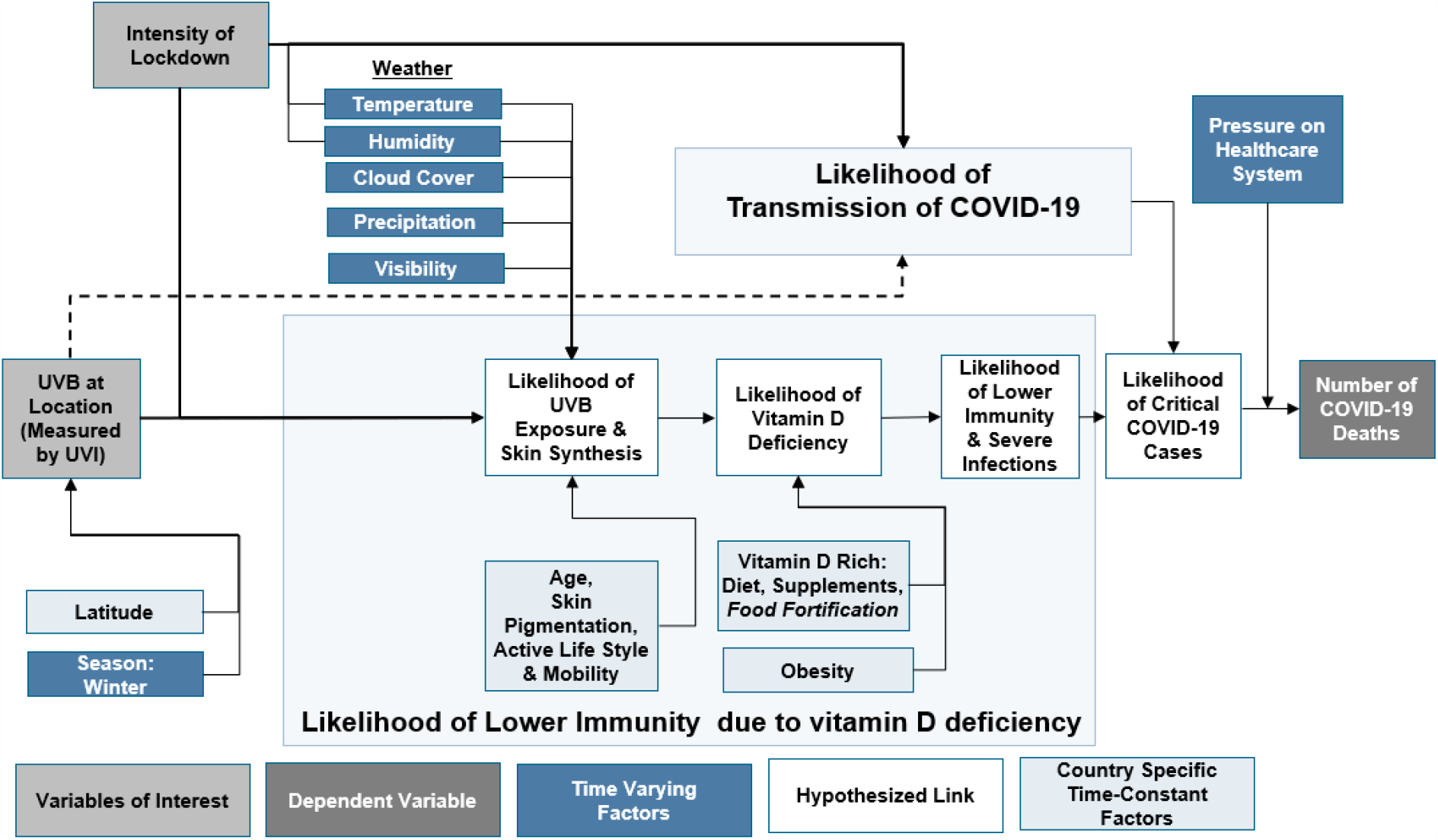
Explanation of the Association of Severity of Lockdown & UVB Radiation with COVID-19 Deaths. Note: We extended the theoretical framework of the protective role of UVB radiation in reducing COVID-19 deaths from our former paper^8^

Although lockdowns might help in reducing the likelihood of transmission of SARS-CoV-2 virus^1^, they potentially reduce the likelihood of UVB radiation exposure, increasing the likelihood of vitamin D deficiency as indicated in Figure 1^8^. Furthermore, emerging studies on COVID-19 indicate the association of vitamin D deficiency with COVID-19 severity and mortality ^4–8^. In light of this emerging evidence, we anticipate that the severity of lockdown and UVB radiation are independently associated with a reduction in the number of COVID-19 deaths. Nevertheless, we anticipate that the increased lockdown severity will be associated with a significant reduction in the protective role of UVI in mitigating COVID-19 deaths due to a reduced likelihood of UVB radiation exposure.

## 3 Data and Methods

### 3.1 Description of Data

In order to empirically estimate the independent protective roles of lockdown, UVB and the mitigating influence of lockdown severity on the protective role of UVB in reducing COVID-19 deaths, we constructed the dataset outlined in Table 1. We collected data covering 108 days from 22 January 2020 until 8 May 2020 across 162 countries of which 142 reported COVID-19 deaths before 8 May 2020 and of which 136 reported more than 20 COVID-19 infections before 8 May 2020. We focus on these 136 countries to ensure that the results are not biased by countries that are at a very early stage of COVID-19 outbreak, which would limit data points concerning COVID-19 deaths. Additionally, we drop the first 20 daily observations of every country after that country reported the first COVID-19 infection to further ensure that the observations do not bias results at the very early stage of the COVID-19 outbreak.

**Table 1:** Summary of Dataset.

|  |  |
| --- | --- |
| Number of countries in the world | 195 |
| Number of countries in our dataset | 162 |
| ... > 0 cumulated COVID-19 deaths before 8 May 2020 | 142 |
| ... > 20 cumulated COVID-19 infections before 8 May 2020 | 136 |
| Covered time-period | 22 January 2020 - 8 May 2020 (108 days) |
| Granularity of data | Daily |
| COVID-19 data source | https://github.com/CSSEGISandData/COVID-19 |
| Weather data source | https://darksky.net/ |
| Lockdown severity data source | https://www.bsg.ox.ac.uk/research/research-projects/coronavirus-government-response-tracker |

The corresponding country-level data consist of the cumulative daily COVID-19 deaths and infections, the daily ultraviolet index (UVI) (closely associated with UVB), and a set of control variables such as daily weather parameters such as precipitation index, cloud index, ozone level, visibility level, humidity level, as well as minimum and maximum temperature. The country-level data also consists of the severity of lockdown enforced by the country’s government. We use the interaction of the lockdown severity with UVI to examine whether higher lockdown severity is associated with a reduction in the protective role of UVI in mitigating COVID-19 deaths.

We present descriptive statistics of the dataset in Table 2. As of 8^th^ of May, the cumulative COVID-19 deaths of these 136 countries were on average 2,020 and the growth rate of COVID-19 deaths on May 8 was on average 2.7% as compared to the average growth rate of COVID-19 deaths across countries and time which was 10.3%. We use cumulative COVID-19 deaths as the main dependent variable to test our hypothesis that lockdowns mitigate the protective role of UVB radiation. Based on the severity of governmental advice and restrictions, there are 4 different severity levels which could decrease skin exposure to UVB radiation ranging from no measure to requiring the population not to leave the house with minimum exceptions. On 8^th^ of May, 15 countries had not implemented a lockdown, 35 countries recommended not to leave the house, 69 required people not to leave the house except for daily exercises, grocery shopping and essential trips, and 17 countries implemented a severe lockdown and required their people not to leave the house with minimal exceptions. UVI is on average 6.84 representing a moderate to high risk of harm from unprotected sun exposure.

**Table 2:**
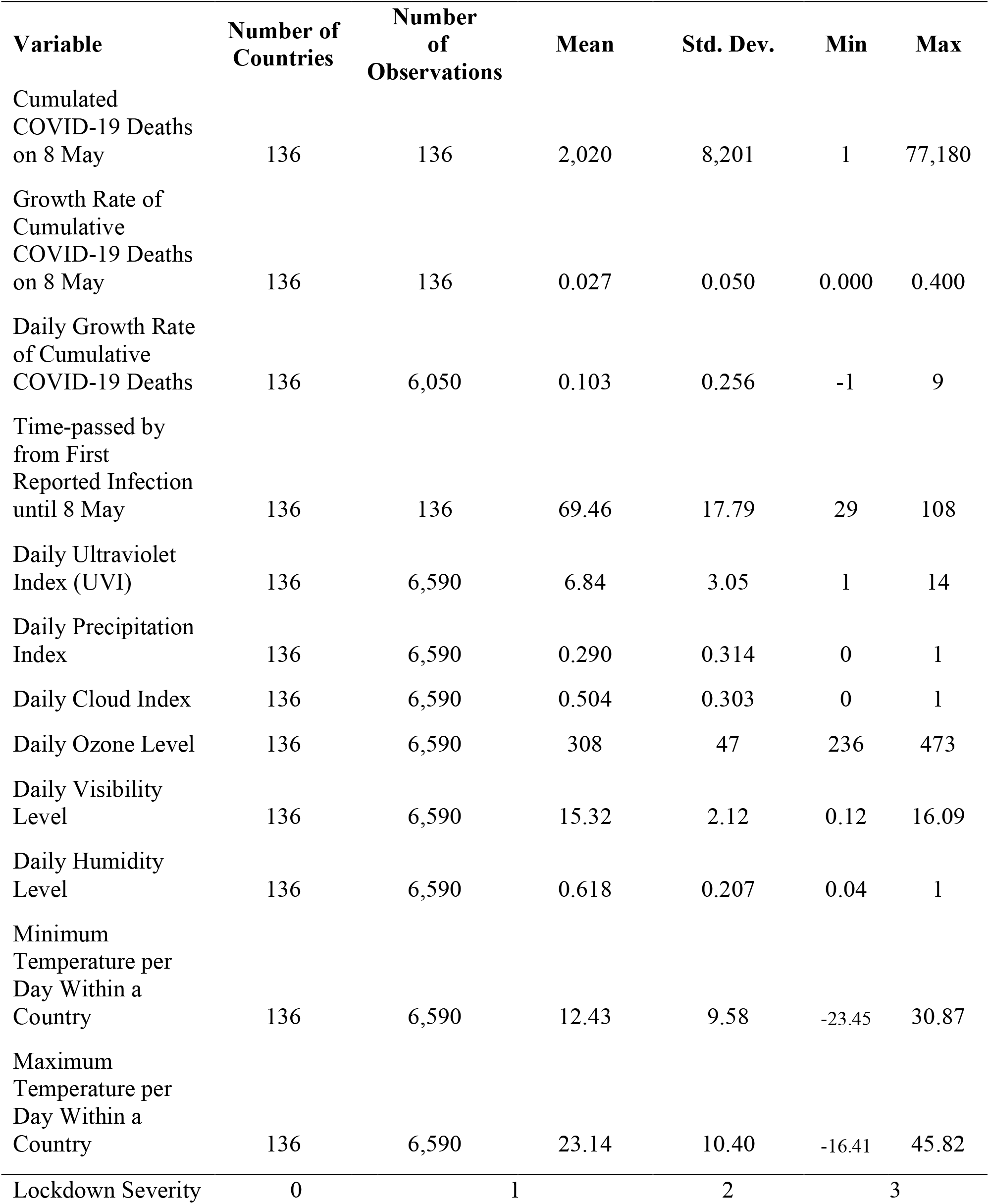
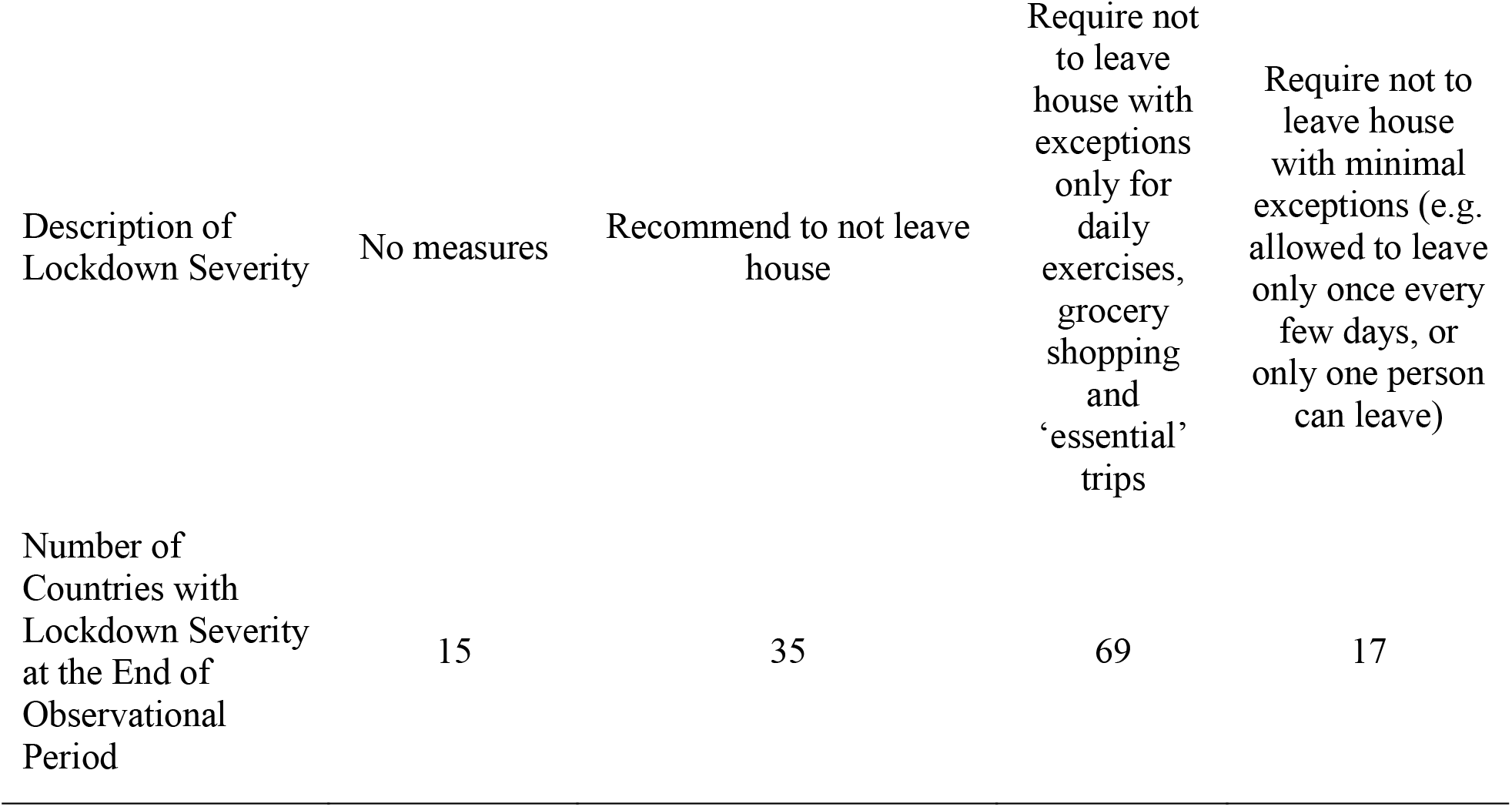
Descriptive Statistics.

### 3.2 Summary of Method

We apply a log-linear fixed-effects model to estimate the mitigating influence of lockdown severity on the association between UVI and growth-rates of COVID-19 deaths. We describe the model and the structural equation which we estimate in the supplementary material building upon Moozhipurath et al. (2020)^8^ as well as Hsiang et al. (2020)^2^.

In order to assess the respective protective roles of lockdown and UVI in mitigating the growth-rates of COVID-19 deaths and subsequently determine whether and which lockdown severity mitigates the protective role of UVI, we estimate three versions of the log-linear fixed-effects model. Model 1 outlines whether a unit increase in the lockdown severity mitigates the association between UVI and the growth-rates of COVID-19. Model 2 and model 3 outline whether a more severe lockdown measure (e.g., level 2 or level 3) mitigates this association more strongly as compared to a less severe lockdown (level 1).

## 4 Results

We estimate the protective role of lockdown severity, UVI and the mitigating influence of lockdown on UVI’s protective role by using the log-linear fixed-effects model. We isolate the mitigating influence of lockdown severity from time-constant country-specific factors (see Figure 1). Further, we use the partialling-out property to isolate the mitigating influence of lockdown severity from all linear as well as some non-linear effects of time-varying factors such as weather and time, which may confound the results.

After controlling for time-constant and time-varying factors, we find that a unit increase in UVI and lockdown severity are independently associated with 1.8 percentage points (p.p) and 7.9 p.p decline in COVID-19 deaths growth rate, indicating their respective protective roles. These declines represent significant percentage reductions in the average growth-rates of COVID-19 deaths of −17% (=−0.018 / 0.103) and −77% (=−0.079/0.103).

However, we find a significant mitigating influence of lockdown severity on the protective role of UVI in reducing the growth-rates of COVID-19 deaths. A unit increase of the lockdown severity weakens the association of UVI in reducing the growth-rates of COVID-19 deaths by −33% (=0.006/−0.018). This decrease represents the average mitigation of a unit increase of the lockdown severity from 0 to 1, 1 to 2, and 2 to 3.

Surprisingly, Model 2 and Model 3 outline that the mitigation effect is mostly associated with lockdown severity of level 1 rather than level 2 or level 3 (stricter lockdowns) as the interaction of lockdown severity of level 2 or 3 with UVI is insignificant. Besides, the lockdown severity of level 1 mitigates the association of UVI and growth-rates of COVID-19 deaths by −95%, (0.018 / −0.019) and practically completely mitigating the association. Finally, all models 1 – 3 outline the significant negative association of both, lockdown severity as well as UVI, with growth-rates of COVID-19 deaths, indicating their protective roles.

We compare two scenarios to illustrate the mitigated protective role of UVI on the cumulative COVID-19 deaths. In scenario 1 UVI’s protective role is not mitigated by lockdown whereas in scenario 2 UVI’s protective role is fully mitigated. In order to relate the mitigated protective role with COVID-19 deaths we take the average number of COVID-19 deaths at the end of the observational period, i.e., 2,020, as cumulative COVID-19 deaths at day 0 as shown in Figure 2. In the full mitigation scenario (Scenario 2), we use the growth-rate of COVID-19 deaths in our sample, i.e., 10.3%. In Scenario 1, where UVI’s role is not mitigated, we use an average growth rate of 8.5% (i.e., 10.3 p.p - 1.8 p.p). Figure 2 outlines that this mitigating influence of lockdown on the protective role of UVI translates into 1,640 or 21% fewer COVID-19 deaths after 14 days.

**Figure 2:**
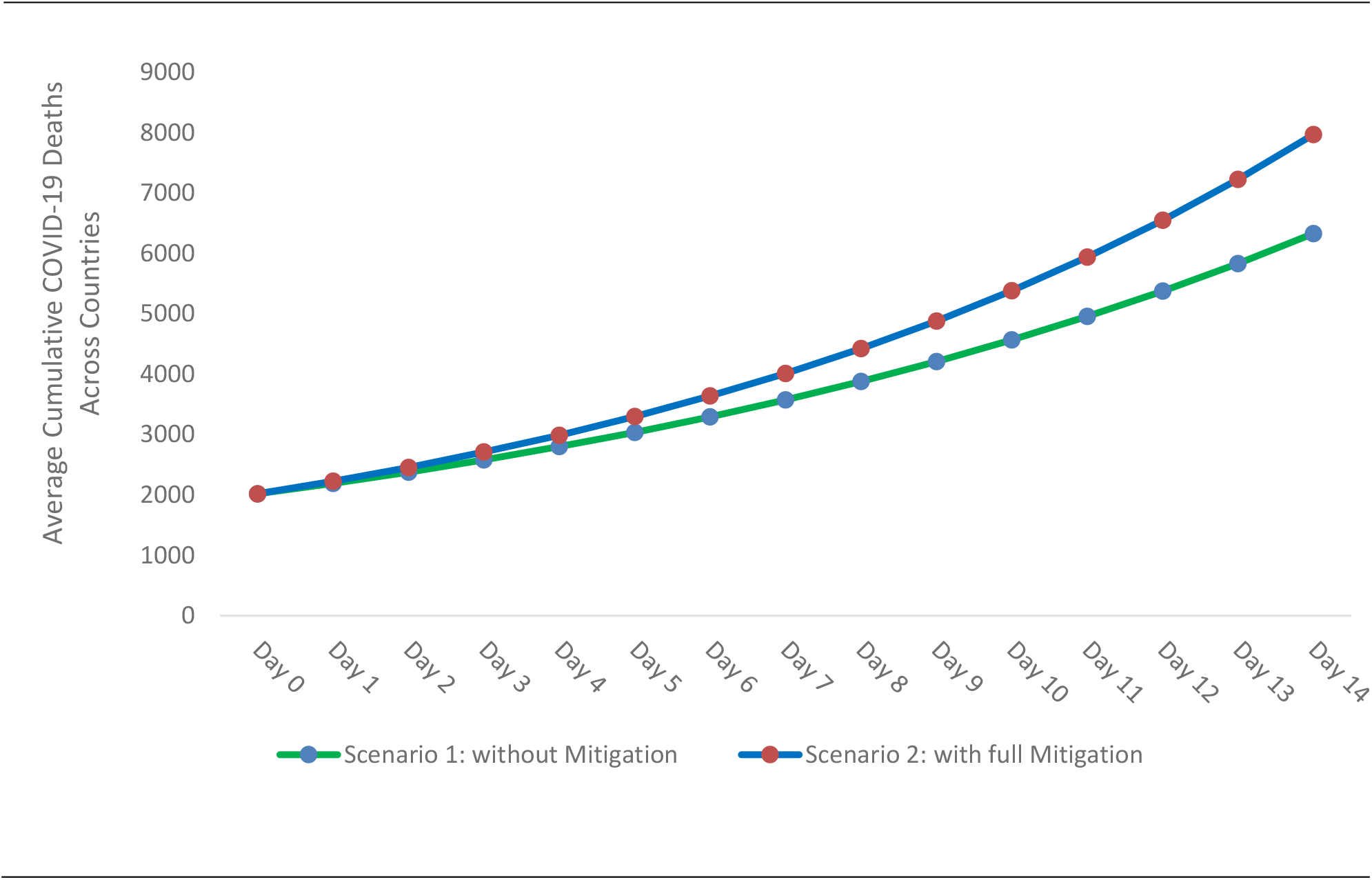
**Comparison of “No Mitigation of UVI’s Protective Role by Lockdown” vs. “Full Mitigation of UVI’s Protective role by Lockdown” on Average Cumulative COVID-19 Deaths Across Countries**

## 5 Discussion

Our empirical results indicate that although large-scale lockdowns are associated with a slowdown in the daily growth rates of COVID-19 deaths, such measures also significantly reduce the protective role of UVB in COVID-19 deaths.

We find that a unit increase in UVI and lockdown severity are independently associated with a decline in COVID-19 deaths growth rate, indicating their respective protective roles. However, the lowest lockdown severity (recommendation not to leave the house) already practically completely mitigates the protective role of UVI in reducing the growth rate of COVID-19 deaths via a reduction of 1.8 percentage points or −95% [p < 0.01]. Our results are consistent across different model specifications.

Our results suggest that lockdowns in conjunction with adequate exposure to UVB radiation might have provided even more substantial health benefits, than lockdowns alone. For example, we estimate that there would be 21% fewer deaths on average with sufficient UVB exposure while people were recommended not to leave their house.

While we acknowledge there may be other confounding factors posing challenges to our analysis, we used advanced statistical methods to account for such factors as much as possible. Even though we anticipate that reduced likelihood of skin synthesis due to lockdown plausibly explain these association, we may not be able to rule out the possibility of other UVB induced mediators – such as nitric oxide ^10,29^.

We follow a macro-level statistical backwards-looking approach which captures the real-life behaviour without making any specific assumptions regarding any epidemiological parameters^2^. Although this macro-level approach is a key strength of the study, the results cannot be interpreted as health guidance, which often comes from clinical studies^2^. Therefore, further clinical studies are needed to establish a causal relationship between UVB induced vitamin D and COVID-19 deaths.

Nations are implementing strict lockdowns to slow down COVID-19 growth. Even though the studies suggest the need for continued interventions^1,2^, in addition to substantial economic and social costs, an unintended consequence of such large-scale interventions is the limited UVB exposure, plausibly increasing the risk of COVID-19 deaths. We require further studies to investigate if COVID-19 deaths can be mitigated by proper social distancing along with sensible exposure to sunlight or via vitamin D intervention. This type of intervention will be desirable from a policy maker’s perspective because of its lower social and economic costs.

Further, nations may consider creating awareness among the population regarding the importance of sensible sunlight exposure while implementing lockdowns. Specifically, nations can look at assisting the vulnerable population who are at a higher risk of vitamin D deficiency – e.g., darker-skinned people living in high latitudes, people with limited mobility or indoor lifestyle (nursing home residents) and vegetarians^8^. We hope that the findings of this study can support policy decision making related to COVID-19 in countries which are currently implementing lockdowns or are considering them.

## Data Availability

The data used in the study are from publicly available sources. Data regarding COVID-19 are obtained on 9th May 2020 from COVID-19 Data Repository by the Center for Systems Science and Engineering (CSSE) at Johns Hopkins University and can be accessed at https://github.com/CSSEGISandData/COVID-19. We will make specific dataset used in this study available for any future research. Interested researchers can contact one of the authors via email to get access to the data.

## 7 Acknowledgements

We would like to acknowledge Bernd Skiera for his immense contribution to this paper and for providing inputs to this paper. We would like to acknowledge Sharath Mandya Krishna, and Rukhshan Ur Rehman for their immense contribution to this paper - for providing inputs and assisting with data collection, data transformation and data engineering. We thank Matthew Little for his inputs and his assistance in review. We would also like to acknowledge Magdalena Ceklarz for her valuable contributions to our paper and the discussions about COVID-19.

## 8 Author Contributions

RKM conceptualized the research idea, conducted literature research and designed theoretical framework. RKM and LK collected the data. LK designed empirical methods and analyzed the data. RKM and LK interpreted the results and wrote the article. RKM and LK reviewed and revised the article.

## 10 Additional Information

Correspondence and requests for materials should be addressed to Rahul Kalippurayil Moozhipurath.

## 11 Data Sharing

The data used in the study are from publicly available sources. Data regarding COVID-19 are obtained on 9^th^ May 2020 from *COVID-19 Data Repository* by the *Center for Systems Science and Engineering (CSSE)* at *Johns Hopkins University* and can be accessed at https://github.com/CSSEGISandData/COVID-19. Data regarding weather is obtained from *Dark Sky* on the 9^th^ May 2020 and can be accessed at https://darksky.net/. Data regarding lockdown severity is obtained from https://www.bsg.ox.ac.uk/research/research-projects/coronavirus-government-response-tracker. We will make specific dataset used in this study available for any future research. Interested researchers can contact one of the authors via email to get access to the data.

## 13 Supplementary Material

### 13.1 Description of Methodology

We apply a fixed-effect log-linear regression model to estimate the effect of UVI on the number of COVID-19 deaths that builds upon Figure 1. The model is closely related to Moozhipurath et al. (2020)^8^ as well as Hsiang et al. (2020)^2^. The major difference to Moozhipurath et al. (2020)^8^ is the introduction of two additional sets of variables. Those additional sets consist of (i) variables representing the lockdown severity and (ii) variables representing the interaction of the lockdown severity and UVI. Thus, we use the following model to explain the number of COVID-19 deaths:

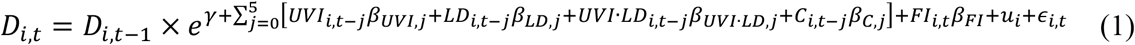

*D*_*i,t*_ represents the cumulative COVID-19 deaths for country *i* at time point *t* (in days) and it is related to the explanatory factors via an exponential growth model on the right-hand side of the equation (1). The exponential growth model flexibly allows for different shapes of the cumulative COVID-19 deaths.

The exponential growth model consists of eight explanatory parts.

1. *γ* represents the daily growth rate of COVID-19 deaths from *D*_*i,t*−1_ to *D*_*i,t*_ that is independent of the factors presented in Figure 1. *γ* covers virus-specific attributes like its basic reproductive rate R_0_ combined with its lethality.
2. *UVI*_*i,t*−*j*_ represents the *UVI* for a country *i* at day *t* lagged by *j* weeks. *β*_*UVI,j*_ reflects the effect of *UVI* lagged by *j* weeks.
3. *LD*_*i,t*−*j*_ represents the lockdown severity *LD* for a country *i* at day *t* lagged by *j* weeks. *β*_*LD,j*_ reflects the effect of *LD* lagged by *j* weeks. *LD* can either consist of one variable lagged by *j* weeks as in Model 1 or it can consist of 2 dummy variables as in model 2 or 3. The first dummy variable of model 2 is equal to one if the lockdown severity is at least equal to 1 whereas the second dummy variable of model 2 is equal to one if the lockdown severity is at least equal to two. The first dummy variable of model 3 is equal to one if the lockdown severity is at least equal to 1 whereas the second dummy variable of model 3 is equal to one if the lockdown severity is at least equal to three. Similarly, *β*_*LD,j*_ can measure the effect of unit increase in the lockdown severity as in model 1. *β*_*LD,j*_ can also measure the effect of a specific lockdown severity as in model 2 and model 3.
4. *UVI* · *LD*_*i,t*−*j*_ represents the interaction of *UVI*_*i,t*−*j*_ and the lockdown severity *LD*_*i,t*−*j*_ for a country *i* at day *t* lagged by *j* weeks. *β*_*UVI*·*LD,j*_ reflects the effect of this interaction lagged by *j* weeks. We either interact *UVI* with variable *LD* for model 1 and we interact *UVI* with the dummy variable for model 2 and model 3. Therefore, *β*_*UVI*·*LD,j*_ reflects the effect of a unit increase of *UVI* lagged by *j* weeks if the lockdown severity increases by one unit as for model 1. *β*_*UVI*·*LD,j*_ also reflects the effect of a unit increase of *UVI* for model 2 and 3 if the lockdown severity is 1 as compared to when the lockdown severity is 0 as well as if the lockdown severity is 2 or 3 as compared to when the lockdown severity is 1.
5. *C*_*i,t*−*j*_ stands for the set of control variables. This set consists of precipitation index, cloud index, ozone level, visibility level, humidity level, as well as minimum and maximum temperature for a country *i* at day *t* lagged by *j* weeks. The vector *β*_*C,j*_ identifies the effect of these control variables lagged by *j* weeks.
6. *FI*_*i,t*_ stands for the time passed by since the first reported COVID-19 infection for a country *i* at day *t* and *β*_*FI*_ identifies the associated effect.
7. *u*_*i*_ represents time-constant country-specific factors influencing the growth rate of cumulative COVID-19 deaths (e.g., diet related effects, population parameters about their activities and demographic composition).
8. *ϵ*_*i,t*_ consists of all the remaining factors that are not identified but also have an effect on the cumulative COVID-19 deaths (i.e., all non-linear differences of growth rates with respect to time and country-specific linear differences of growth rates with respect to time. They could be caused by a decreasing number of people who could potentially become infected or contagious, lockdowns in a country over time, mutation of the virus in a country over time, systematic false-reports of the dependent variable).

An appropriate transformation outlined in (Moozhipurath et. al 2020) results in the estimable equation

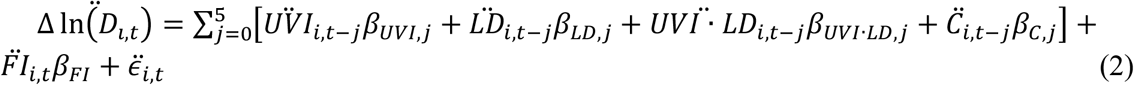

*γ* and *u*_*i*_ do not appear in the equation anymore and a linear regression can identify all other coefficients. The summation of the coefficients *β* for each variable measures the long-term effect of a permanent unit change of the respective variable and outlines by how many percentage points the growth-rate of COVID-19 deaths changes. If 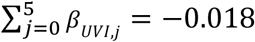, then a permanent unit increase of UVI is associates with a 1.8 percentage points decline in the growth rates of COVID-19 deaths.

Equation (2) also shows why we can only use those observations where cumulative COVID-19 deaths are larger than zero. This condition explains the difference between the 6,590 observations of 136 countries in Table 2 and the 6,049 observations of 136 countries in Table 3. We listed the number of observations per country we used in the analysis in Table S3.

**Table 3:** Results of Log-Linear Fixed-Effects Model.

|  | Model 1 | Model 2 | Model 3 |
| --- | --- | --- | --- |
| Dependent Variables | COVID-19 Deaths | COVID-19 Deaths | COVID-19 Deaths |
| UVI | -0.018*** (12.60) | -0.019*** (11.47) | -0.019*** (10.75) |
| LD | -0.079*** (18.71) |  |  |
| LD x UVI | 0.006** (8.51) |  |  |
| LD severity 1 |  | -0.167** (9.74) |  |
| LD severity 1 x UVI |  | 0.018** (7.32) |  |
| LD severity 2 or 3 |  | 0.003 (0.01) |  |
| LD severity 2 or 3 x UVI |  | -0.006 (1.20) |  |
| LD severity 1 or 2 |  |  | -0.172*** (11.88) |
| LD severity 1 or 2 x UVI |  |  | 0.015* (5.87) |
| LD severity 3 |  |  | 0.020 (0.27) |
| LD severity 3 x UVI |  |  | -0.003 (0.50) |
| **Control Variables** |  |  |  |
| Time Trend | Linear | Linear | Linear |
| Country Fixed Effects | Yes | Yes | Yes |
| Precipitation index | Yes | Yes | Yes |
| Cloud index | Yes | Yes | Yes |
| Ozone level | Yes | Yes | Yes |
| Visibility Level | Yes | Yes | Yes |
| Humidity level | Yes | Yes | Yes |
| Temperature (min and max) | Yes | Yes | Yes |
| Number of Estimates | 61 (+ 136 FE) | 73 (+ 136 FE) | 73 (+ 136 FE) |
| Number of Observations | 6,049 | 6,049 | 6,049 |
| Number of Countries | 136 | 136 | 136 |
| R-squared Within | 16.86% | 17.20% | 17.07% |
| Note: +: $p < 0.10$, *: $p < 0.05$, **: $p < 0.01$. F-statistic for long-run coefficients in parentheses. | | | |

### 13.2 Model Selection

We estimated different versions of model which varied in the number of weekly lags. We decided to choose a model with 5 weekly lags because we could not find major differences with respect to the estimated coefficients when increasing the number of lags and a more parsimonious is favorable.

### 13.3 Robustness Checks

In order to assess the robustness of our results of the primary model - Model 1 - we isolate the mitigating influence of lockdown severity on the association of UVI and growth-rates of COVID-19 deaths from time trends in flexible ways. Models 4 – 9 in Table S1 and Table S2 isolate our findings from linear, square and exponential time trends which may be similar across countries or even country-specific. Overall, we find consistent results across model specifications. A permanent unit increase of UVI and a permanent unit increase in lockdown severity are independently associated with a −1.2 – −1.8 and −7.6 – −12.0 percentage points reduction in growth-rates of COVID-19 deaths, respectively, according to different model specifications. Results also indicate that a permanent unit increase in lockdown severity weakens the association of UVI in reducing the growth-rates of COVID-19 deaths by 0.4 –0.6 percentage points.

**Table S1:** Robustness Checks with Random-Effects Model.

|  | Model 4 | Model 5 | Model 6 |
| --- | --- | --- | --- |
| Dependent Variables | COVID-19 Deaths | COVID-19 Deaths | COVID-19 Deaths |
| UVI | -0.018*** (12.96) | -0.012* (5.35) | -0.014* (6.72) |
| LD | -0.076*** (16.36) | -0.107*** (29.19) | -0.120*** (21.05) |
| LD x UVI | 0.006** (8.87) | 0.004 (2.60) | 0.005 (2.74) |
| **Control Variables** |  |  |  |
| Time Trend | Linear and Square | Country-specific Linear | Country-specific Linear and Square |
| Country Fixed Effects | Yes | Yes | Yes |
| Precipitation index | Yes | Yes | Yes |
| Cloud index | Yes | Yes | Yes |
| Ozone level | Yes | Yes | Yes |
| Visibility Level | Yes | Yes | Yes |
| Humidity level | Yes | Yes | Yes |
| Temperature (min and max) | Yes | Yes | Yes |
| Number of Estimates | 62 (+ 136 FE) | 60 (+ 136 FE + 136 TCSE) | 60 (+ 136 FE + 272 TCSE) |
| Number of Observations | 6,049 | 6,049 | 6,049 |
| Number of Countries | 136 | 136 | 136 |
| R-squared Within | 16.90% | 20.85% | 24.68% |

Note: +: $p < 0.10$ , \*: $p < 0.05$ , \*\*: $p < 0.01$ . F-statistic for long-run coefficient in parentheses. TCSE stands for time country-specific effects.

**Table S2:** Robustness Checks with Random-Effects Model.

|  | **Model 7** | **Model 8** | **Model 9** |
| --- | --- | --- | --- |
| Dependent Variables | COVID-19 Deaths | COVID-19 Deaths | COVID-19 Deaths |
| UVI | -0.018*** (12.64) | -0.017*** (11.90) | -0.014** (6.91) |
| LD | -0.078*** (18.45) | -0.080*** (18.12) | -0.121*** (21.20) |
| LD x UVI | 0.006** (8.52) | 0.006** (8.48) | 0.005+ (2.83) |
| **Control Variables** |  |  |  |
| Time Trend | Linear and Exponential | Linear and Country-specific Exponential | Country-specific Linear, Square and Exponential |
| Country Fixed Effects | Yes | Yes | Yes |
| Precipitation index | Yes | Yes | Yes |
| Cloud index | Yes | Yes | Yes |
| Ozone level | Yes | Yes | Yes |
| Visibility Level | Yes | Yes | Yes |
| Humidity level | Yes | Yes | Yes |
| Temperature (min and max) | Yes | Yes | Yes |
| Number of Estimates | 62 (+ 136 FE) | 61 (+ 136 FE + 136 TSCE) | 60 (+ 136 FE + 408 TSCE) |
| Number of Observations | 6,049 | 6,049 | 6,049 |
| Number of Countries | 136 | 136 | 136 |
| R-squared Within | 16.86% | 17.16% | 25.29% |

Note: +: $p < 0.10$ , \*: $p < 0.05$ , \*\*: $p < 0.01$ . F-statistic for long-run coefficient in parentheses. TCSE stands for time country-specific effects.

**Table S3:** Number of Observations (Obs,) of Countries Used in Analysis.

| Country | Obs. | Country | Obs. | Country | Obs. | Country | Obs. |
| --- | --- | --- | --- | --- | --- | --- | --- |
| Afghanistan | 47 | Czechia | 47 | Kazakhstan | 38 | Russia | 50 |
| Albania | 42 | Denmark | 53 | Kenya | 38 | San Marino | 53 |
| Algeria | 55 | Djibouti | 28 | Korea, South | 73 | Saudi Arabia | 45 |
| Andorra | 47 | Dominican Republic | 50 | Kosovo | 25 | Senegal | 37 |
| Angola | 31 | Ecuador | 50 | Kuwait | 34 | Serbia | 45 |
| Argentina | 48 | Egypt | 61 | Kyrgyzstan | 33 | Sierra Leone | 15 |
| Australia | 68 | El Salvador | 32 | Latvia | 32 | Singapore | 48 |
| Austria | 55 | Estonia | 44 | Lebanon | 59 | Slovakia | 37 |
| Azerbaijan | 50 | Eswatini | 22 | Liberia | 34 | Slovenia | 46 |
| Bahrain | 53 | Ethiopia | 33 | Libya | 27 | Somalia | 30 |
| Bangladesh | 43 | Finland | 48 | Lithuania | 48 | South Africa | 42 |
| Barbados | 33 | France | 73 | Luxembourg | 51 | Spain | 66 |
| Belarus | 38 | Gabon | 37 | Malawi | 18 | Sri Lanka | 41 |
| Belgium | 58 | Georgia | 34 | Malaysia | 52 | Sudan | 38 |
| Benin | 32 | Germany | 60 | Mali | 26 | Sweden | 58 |
| Bolivia | 40 | Ghana | 37 | Mauritius | 33 | Switzerland | 55 |
| Bosnia and Herzegovina | 46 | Greece | 54 | Mexico | 50 | Syria | 29 |
| Botswana | 21 | Guatemala | 37 | Moldova | 43 | Taiwan* | 73 |
| Brazil | 52 | Guinea | 23 | Morocco | 49 | Tanzania | 35 |
| Brunei | 41 | Guyana | 39 | Netherlands | 53 | Thailand | 68 |
| Bulgaria | 43 | Haiti | 31 | New Zealand | 40 | Togo | 42 |
| Burkina Faso | 41 | Honduras | 40 | Niger | 31 | Trinidad and Tobago | 37 |
| Cameroon | 44 | Hungary | 47 | Nigeria | 46 | Tunisia | 47 |
| Canada | 60 | Iceland | 50 | Norway | 54 | Turkey | 40 |
| Chad | 10 | India | 58 | Oman | 38 | US | 69 |
| Chile | 47 | Indonesia | 49 | Pakistan | 50 | Ukraine | 48 |
| China | 73 | Iran | 61 | Panama | 41 | United Arab Emirates | 49 |
| Colombia | 45 | Iraq | 56 | Paraguay | 43 | United Kingdom | 63 |
| Congo (Kinshasa) | 40 | Ireland | 51 | Peru | 45 | Uruguay | 37 |
| Costa Rica | 45 | Israel | 48 | Philippines | 73 | Uzbekistan | 36 |
| Cote d'Ivoire | 40 | Italy | 73 | Poland | 47 | Venezuela | 37 |
| Croatia | 50 | Jamaica | 40 | Portugal | 49 | Yemen | 8 |
| Cuba | 39 | Japan | 73 | Qatar | 41 | Zambia | 33 |
| Cyprus | 42 | Jordan | 42 | Romania | 47 | Zimbabwe | 31 |
| Total Number of Observations |  |  |  | 6,049 |  |  |  |

## References

1. Flaxman, S. et al. Estimating the effects of non-pharmaceutical interventions on COVID-19 in Europe. Nature 1–8 (2020) doi:10.1038/s41586-020-2405-7.

2. Hsiang, S. et al. The effect of large-scale anti-contagion policies on the COVID-19 pandemic. Nature 1–9 (2020) pdoi:10.1038/s41586-020-2404-8.

3. Chinazzi, M. et al. The effect of travel restrictions on the spread of the 2019 novel coronavirus (COVID-19) outbreak. Science eaba9757 (2020) doi:10.1126/science.aba9757.

4. Grant, W. B. et al. Evidence that Vitamin D Supplementation Could Reduce Risk of Influenza and COVID-19 Infections and Deaths. Nutrients 12, 988 (2020).

5. Watkins J. Preventing a covid-19 pandemic. BMJ 2020;368:m810.published online February 2020.

6. Panarese, A. & Shahini, E. Letter: Covid-19, and vitamin D. Aliment. Pharmacol. Ther. 51, 993–995 (2020).

7. Lanham-New, S. A. et al. Vitamin D and SARS-CoV-2 virus/COVID-19 disease. BMJ Nutr. Prev. Health (2020) doi:10.1136/bmjnph-2020-000089.

8. Kalippurayil Moozhipurath, R., Kraft, L. & Skiera, B. Evidence of Protective Role of Ultraviolet-B (UVB) Radiation in Reducing COVID-19 Deaths. https://papers.ssrn.com/abstract=3586555 (2020) doi:10.2139/ssrn.3586555.

9. Kraemer, M. U. G. et al. The effect of human mobility and control measures on the COVID-19 epidemic in China. 6 (2020).

10. Hart, P. H., Gorman, S. & Finlay-Jones, J. J. Modulation of the immune system by UV radiation: more than just the effects of vitamin D? Nat. Rev. Immunol. 11, 584–596 (2011).

11. Bodiwala, D. et al. Prostate cancer risk and exposure to ultraviolet radiation: further support for the protective effect of sunlight. Cancer Lett. 192, 145–149 (2003).

12. Grant, W. B. An estimate of premature cancer mortality in the US due to inadequate doses of solar ultraviolet-B radiation. Cancer 94, 1867–1875 (2002).

13. Grant, W. B. An ecologic study of the role of solar UV-B radiation in reducing the risk of cancer using cancer mortality data, dietary supply data, and latitude for European countries. in Biologic Effects of Light 2001 267–276 (Springer, 2002).

14. Rostand, S. G. Ultraviolet light may contribute to geographic and racial blood pressure differences. Hypertension 30, 150–156 (1997).

15. Holick, M. F. Vitamin D deficiency. N. Engl. J. Med. 357, 266–281 (2007).

16. Zittermann, A. Vitamin D in preventive medicine: are we ignoring the evidence? Br. J. Nutr. 89, 552–572 (2003).

17. Tangpricha, V., Pearce, E. N., Chen, T. C. & Holick, M. F. Vitamin D insufficiency among free-living healthy young adults. Am. J. Med. 112, 659–662 (2002).

18. Semba, R. D., Garrett, E., Johnson, B. A., Guralnik, J. M. & Fried, L. P. Vitamin D deficiency among older women with and without disability. Am. J. Clin. Nutr. 72, 1529– 1534 (2000).

19. Wortsman, J., Matsuoka, L. Y., Chen, T. C., Lu, Z. & Holick, M. F. Decreased bioavailability of vitamin D in obesity. Am. J. Clin. Nutr. 72, 690–693 (2000).

20. Bouillon, R. et al. Skeletal and extraskeletal actions of vitamin D: current evidence and outstanding questions. Endocr. Rev. 40, 1109–1151 (2019).

21. White, J. H. Vitamin D signaling, infectious diseases, and regulation of innate immunity. Infect. Immun. 76, 3837–3843 (2008).

22. Liu, P. T. et al. Toll-like receptor triggering of a vitamin D-mediated human antimicrobial response. Science 311, 1770–1773 (2006).

23. Martineau, A. R. et al. Vitamin D supplementation to prevent acute respiratory tract infections: systematic review and meta-analysis of individual participant data. BMJ 356, i6583 (2017).

24. Martineau, A. R. et al. High-dose vitamin D3 during intensive-phase antimicrobial treatment of pulmonary tuberculosis: a double-blind randomised controlled trial. The Lancet 377, 242–250 (2011).

25. M Perron, R. & Lee, P. Efficacy of high-dose vitamin D supplementation in the critically ill patients. Inflamm. Allergy-Drug Targets 12, 273–281 (2013).

26. Wang, J., Tang, K., Feng, K. & Lv, W. High Temperature and High Humidity Reduce the Transmission of COVID-19. https://papers.ssrn.com/abstract=3551767 (2020) doi:10.2139/ssrn.3551767.

27. Sagripanti, J.-L. & Lytle, C. D. Inactivation of Influenza Virus by Solar Radiation. Photochem. Photobiol. 83, 1278–1282 (2007).

28. Skutsch, M. et al. The association of UV with rates of COVID-19 transmission and deaths in Mexico: the possible mediating role of vitamin D. medRxiv 2020.05.25.20112805 (2020) doi:10.1101/2020.05.25.20112805.

29. Deliconstantinos, G., Villiotou, V. & Stravrides, J. C. Release by ultraviolet B (u.v.B) radiation of nitric oxide (NO) from human keratinocytes: a potential role for nitric oxide in erythema production. Br. J. Pharmacol. 114, 1257–1265 (1995).

